# Differential Serum Levels of CACNA1C, Circadian Rhythm and Stress Response Molecules in Subjects with Bipolar Disorder: Associations with Genetic and Clinical Factors

**DOI:** 10.1101/2024.04.11.24305678

**Authors:** Obie Allen, Brandon J. Coombes, Vanessa Pazdernik, Barbara Gisabella, Joshua Hartley, Joanna M. Biernacka, Mark A. Frye, Matej Markota, Harry Pantazopoulos

**Author notes:** **Address for correspondence:** Harry Pantazopoulos, PhD, Department of Psychiatry and Human Behavior, University of Mississippi School of Medicine, TRC 407, 2500 North State St, Jackson, MS 39216, USA.

## Abstract

**Background:** Many patients with bipolar disorder (BD) do not respond to or have difficulties tolerating lithium and/or other mood stabilizing agents. There is a need for personalized treatments based on biomarkers in guiding treatment options. The calcium voltage-gated channel CACNA1C is a promising candidate for developing personalized treatments. CACNA1C is implicated in BD by genome-wide association studies and several lines of evidence suggest that targeting L-type calcium channels could be an effective treatment strategy. However, before such individualized treatments can be pursued, biomarkers predicting treatment response need to be developed.

**Methods:** As a first step in testing the hypothesis that CACNA1C genotype is associated with serum levels of CACNA1C, we conducted ELISA measures on serum samples from 100 subjects with BD and 100 control subjects.

**Results:** We observed significantly higher CACNA1C (p<0.01) protein levels in subjects with BD. The risk SNP (rs11062170) showed functional significance as subjects homozygous for the risk allele (CC) had significantly greater CACNA1C protein levels compared to subjects with one (p=0.013) or no copies (p=0.009). We observed higher somatostatin (SST) (p<0.003) protein levels and lower levels of the clock protein ARTNL (p<0.03) and stress signaling factor corticotrophin releasing hormone (CRH) (p<0.001) in BD. SST and PER2 protein levels were associated with both alcohol dependence and lithium response.

**Conclusions:** Our findings represent the first evidence for increased serum levels of CACNA1C in BD. Along with altered levels of SST, ARNTL, and CRH our findings suggest CACNA1C is associated with circadian rhythm and stress response disturbances in BD.

## Introduction

Bipolar disorder (BD) is a debilitating disease that affects approximately 2.5% of people in the U.S (1), but its etiology and the therapeutic mechanisms of current pharmacological therapies used to treat its symptoms remain unclear. Treatment resistance remains a significant concern in everyday clinical practice of treating patients with BD. Lithium represents the most effective, widely used treatment for BD (2, 3). However, a significant proportion of patients do not respond to lithium therapy or to other commonly used treatments (4, 5), which remains a significant concern in clinical practice and points to the need for personalized treatments based on genetic and biological factors. CACNA1C, which encodes the L-type calcium channel cav1.2, is one of the most strongly implicated genetic factors in genome-wide association studies (GWAS) of BD (6–12) and represents a promising factor for developing personalized treatments. A recent GWAS identified enrichment of polygenic risk factors for targets of calcium channel blockers, including the L-type calcium channel blocker isradipine (13). Several factors associated with CACNA1C, including stress response signaling and circadian rhythm regulation, may be involved in BD symptoms and treatment response (14–20). The L-type calcium channel blocker nimodipine has been reported to be effective in treating both manic and depressive symptoms (14–16). In a group of patients with mood disorders who had low levels of the neurotransmitter somatostatin (SST) in cerebrospinal fluid samples, patients who demonstrated positive response to the L-type calcium channel blocker nimodipine, showed increases in SST cerebrospinal fluid levels after treatment (14), suggesting that therapeutic effects of L-type calcium channel blockers may occur in part through restoration of SST levels. The rs1006737 genetic polymorphism for CACNA1C that was implicated in BD has been shown to result in increased gene expression and increased L-type calcium channel current in cultured human neurons (21), providing further support that L-type calcium channel blockade may be a corrective treatment for genetic effects impacting SST in BD. SST is involved in regulating stress response, anxiety and depression (22–24). Diurnal rhythm alteration through altered light-dark cycles increases anxiety and corticotrophin releasing hormone (CRH) expression in part through altered SST signaling (25). Decreased brain SST expression has been reported in several studies of people with mood disorders as well as in preclinical models of depression (26, 27). Furthermore, altered diurnal rhythm of SST expression was reported in the amygdala of subjects with BD (28). In addition, rats susceptible to chronic mild stress display increased serum levels of SST (29), suggesting SST may represent a peripheral marker for predicting susceptibility for depression. In addition to SST, the stress response molecule CRH may be involved in predicting lithium response. Lithium was reported to increase CRH levels in dexamethasone stress response tests in people with depression who responded to lithium therapy (17–19). Decreased serum CRH in people with BD has also been reported (30).

Circadian rhythm alterations may also be associated with CACNA1C dysfunction and treatment response. For example, CACNA1C genotype predicts the degree of circadian rhythm entrainment of fibroblasts from subjects with BD (12), as well as response to lithium (20). Evidence that CACNA1C is involved in re-setting circadian rhythmicity in the suprachiasmatic nucleus in response to light provides additional support for its role in circadian rhythm regulation (10). Collectively, peripheral measures of molecules such as CACNA1C, CRH, SST and clock molecules may allow for development of blood biomarkers for people with BD and may assist in identifying poor response to lithium as well as people who may benefit from calcium channel blocker and chronobiology based therapeutic strategies. However, information regarding CACNA1C protein levels together with stress signaling and clock molecules in people with BD and how this correlates with genotype and treatment response is lacking, limiting the ability to develop patient specific therapies. We tested the hypothesis that CACNA1C serum protein levels are increased in subjects with BD, along with altered protein levels of clock molecules and stress signaling proteins. Furthermore, we tested the hypothesis that the CACNA1C rs11062170 BD risk allele is also associated with increased CACNA1C protein levels. Finally, we tested whether serum protein levels are correlated with relevant clinical measures including lithium treatment response, psychosis, suicide attempts, mania, hypomania, depression, alcohol dependence, insomnia, and chronotype in subjects with BD.

## Methods

### Human Subjects

Frozen serum samples from 100 self-reported White subjects with BD from Mayo Clinic were randomly selected from the Mayo Clinic Bipolar Disorder Biobank (MCBDB) (31) which has over 2000 cases with BD enrolled and genotyped. From the Mayo Clinic Biobank (MCB) (32), 100 control subjects were matched to cases on age (±5 years), sex, self-reported race, and time of blood draw (±2 hours) and were used for protein serum analysis (Suppl. Table 1). The MCBDB (31, 33) and MCB (32) were both previously described in detail. BD patients were required to have Diagnostic and Statistical Manual of Mental Disorders IV-TR diagnosis of BD-I or BD-II as determined by using the Structured Clinical Interview for DSM-IV (SCID) (34). Potential controls with International Classification of Disease-9 (ICD-9) and/or ICD-10 codes for bipolar disorder and schizophrenia; as well as participants reporting family history of BD or schizophrenia were excluded. This study was approved by the Institutional Review Board of the Mayo Clinic.

### Genotyping and Clinical Measures

Samples from the MCB, comprising 52,865 individuals with adequate sample and consent, and the MCBDB cases recruited at Mayo Clinic (N=1,149) were sequenced at the Regeneron Genetics Center (RGC) with genotype-by-sequencing. The CACNA1C variant rs11062170 was one of the variants sequenced as part of this process and passed quality control filters as has been previously described for the sample (33). The CACNA1C genotypes rs1062170 was provided by both biobanks. Clinical measures at study enrollment for cases were obtained from standardized assessement forms by the MCBDB (see Supplemental Materials). Lithium response was retrospectively assessed using the Alda A score (35).

### ELISA Protein Quantification

CACNA1C, SST, CRH, ARNTL, and PER2 protein levels were measured by enzyme linked immunosorbent assay (ELISA). Commercially available ELISA kits from Abbexa LLC, Sugar Land, TX, were used for quantification of serum levels of human CACNA1C (cat# abx504529, lot# E2205732J), human SST (cat# abx153136, lot# E2205263F), human ARNTL (cat# abx258558, lot# E2210333X), human PER2 (cat# abx382155, lot# E2212916L) and human CRH (cat# abx151177, lot# E2211161A). ELISA plates were measured with an ELISA reader (INFINITE M PLEX, model # INFINTE 200 PRO, TECAN Ltd., Switzerland) and analyzed using Magellan V7.2 SP (TECAN Ltd., Switzerland).

### Statistical Analysis

Descriptive statistics were summarized as frequency and percent or median and interquartile range (IQR) for categorical or continuous variables, respectively. Case-control differences in serum protein levels of CACNA1C, SST, CRH, ARNTL and PER2 were assessed using paired *t*-tests. In addition, plots of serum level by venipuncture time overlaid with each diagnosis group were used to visualize associations of serum levels with time of day. We then tested whether the known CACNA1C risk allele rs11062170 is associated with serum protein levels in CACNA1C among BD and control participants, separately, using linear regression models adjusting for time of day of venipuncture (continuous linear effect), age, and sex.

Secondary analyses were performed to determine whether serum protein levels were also associated with clinical differences within the cases after adjusting for the same set of covariates. Estimates and 95% confidence intervals are reported.

False discovery rate was controlled by adjusting p-values via Benjamini & Hochberg step-up procedure (36, 37). All analyses were conducted using R Statistical Software (v4.2.2; R Core Team 2022) and the multtest R package for false discovery rate control (38, 39).

## Results

### Increased CACNA1C Serum Protein Levels in Subjects with BD

We observed significantly higher CACNA1C serum protein levels in subjects with BD compared to matched control subjects (median [IQR] paired difference=0.024 [−0.007, 0.039]) (p<0.001) (Fig. 1A; Supp Table 1). Furthermore, subjects with BD with two copies of the rs11062170 CC risk allele had significantly greater CACNA1C protein levels (median=0.395 [IQR, 0.391, 0.422]) compared to subjects with BD with one (median=0.392 [IQR, 0.388, 0.393]; p=0.013) or no copies (median=0.391 [IQR, 0.387, 0.395]; p=0.009) of the risk allele (Fig. 1B). In comparison, control subjects with the rs11062170 risk allele did not show differences in CACNA1C protein levels (p=0.145; Fig. 1C).

**Figure 1.**
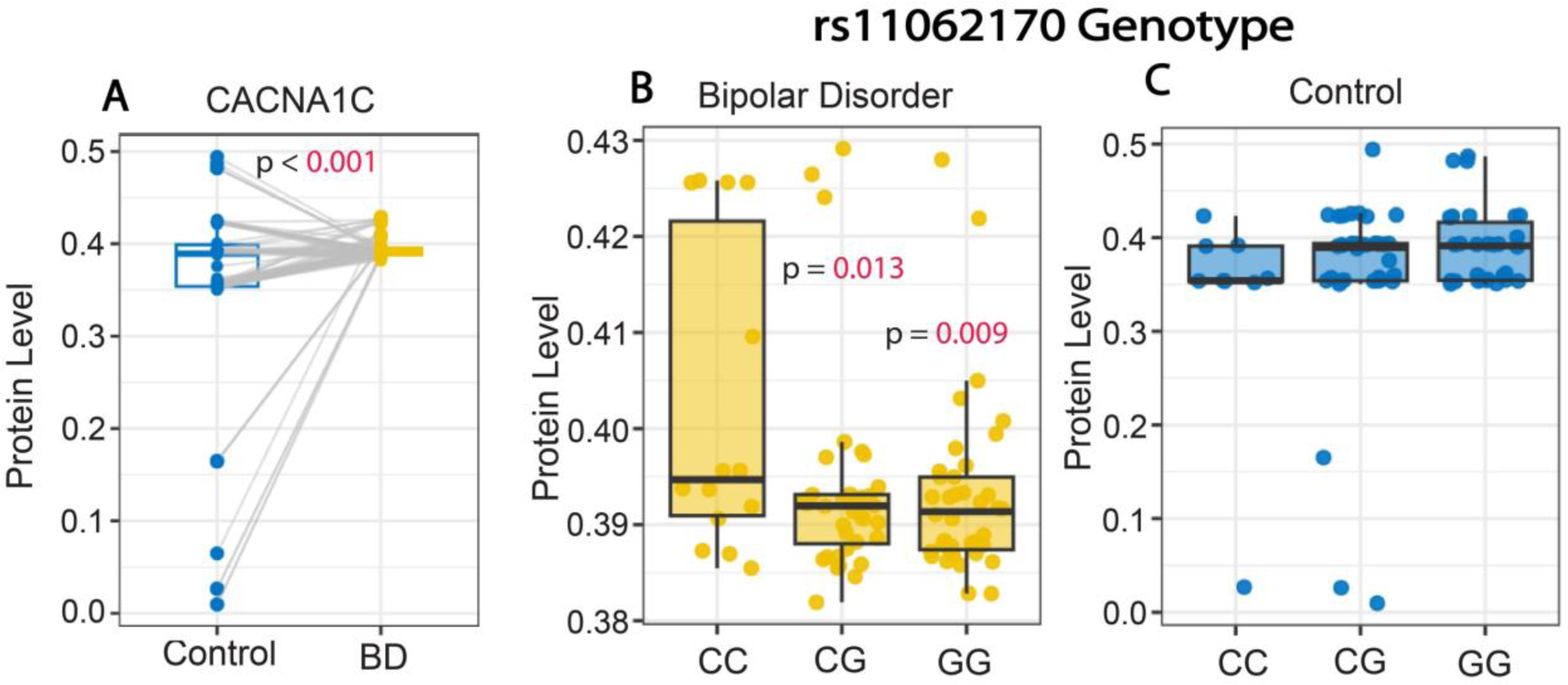
Increased CACNA1C serum protein levels in subjects with bipolar disorder. (A) Whisker plot depicting significantly greater CACNA1C protein levels in serum samples from subjects with BD compared to control subjects. Subjects with BD with two copies of the rs1006737 CC risk allele displayed significantly greater CACNA1C serum protein levels compared to subjects with BD with one or no copies of the CC allele (B). In comparison, no association was observed with the rs1006737 risk allele and CACNA1C protein levels in control subjects (C). Protein values represent ng/ml.

### Altered Serum Levels of Stress Signaling and Circadian Clock Proteins in Subjects with BD

Subjects with BD had significantly higher serum protein levels of SST and lower levels of CRH in comparison to control subjects (median [IQR] paired difference=1.3 [−1.3, 4.7] and –2.4 [−3.6, −1.1], respectively) (both p<0.001) (Fig. 2A&B; Supp Table 1). The circadian clock protein ARNTL displayed significantly lower levels in the serum of subjects with BD compared to control subjects (difference=-0.025 [−0.138, 0.061]; p=0.029), whereas levels of the circadian clock protein PER2 were not statistically different (p=0.103) (Fig. 2C&D; Supp Table 1).

**Figure 2.**
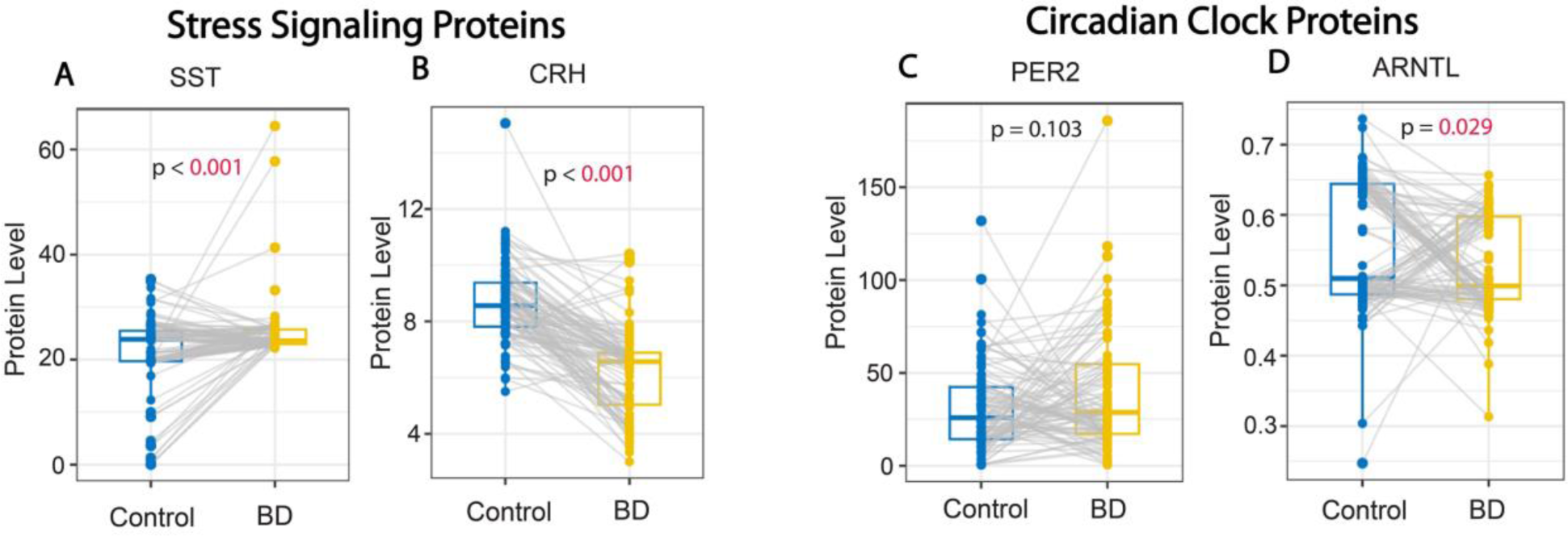
Altered levels of stress signaling and circadian clock proteins in subjects with bipolar disorder. (A) Whisker plots depicting significantly greater SST serum protein levels (A), and lower serum protein levels of CRH (B) in subjects with BD compared to control subjects. No difference was detected for serum levels of the core clock protein PER2 (C) whereas levels of the core clock protein ARNTL were significantly lower in subjects with BD (D). Protein values represent pg/ml for SST and CRH and ng/ml for PER2 and ANRTL.

### Time of Day Alterations in Protein Levels in Subjects with BD

Qualitative analysis of time-of-day alterations in serum levels revealed decreased ARNTL protein and increased SST protein in subjects with BD were most pronounced between 10 am-1 pm (Fig. 3). In comparison CRH protein levels were consistently decreased throughout the day (Fig. 3). PER2 and CACNA1C protein levels were largely consistent across the day (Fig. 3).

**Figure 3.**
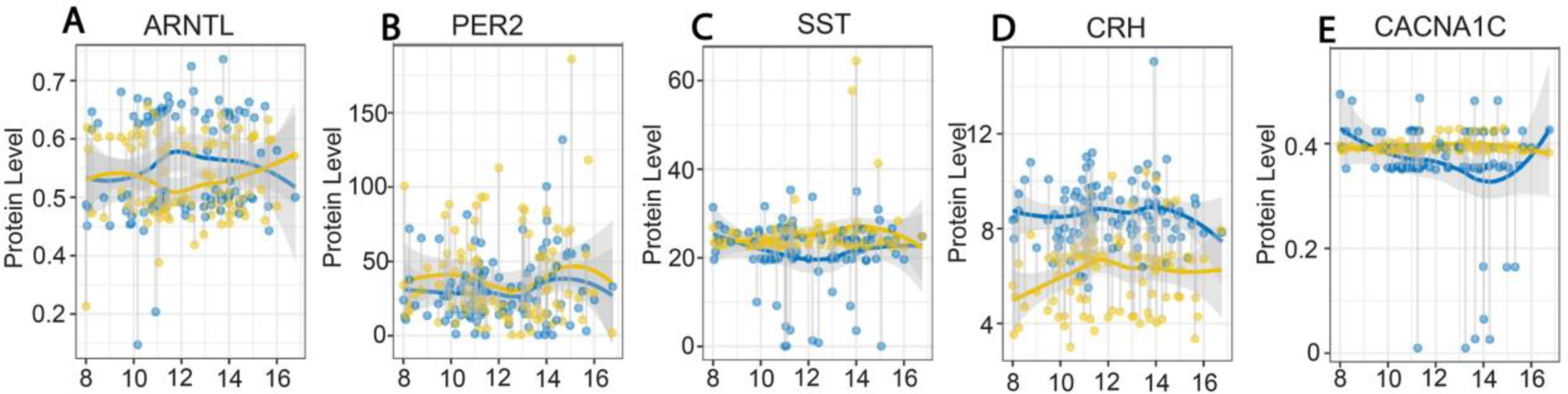
Fluctuations in serum protein levels across the day in subjects with bipolar disorder and control subjects. Plots depicting serum protein levels from samples collected between 8 AM and 5 PM in control subjects (blue) and subjects with BD (gold). ARNTL protein levels were lowest between 10 AM-2 PM in subjects with BD (A) whereas PER2 levels displayed minimal differences across the day compared to control subjects (B). SST protein levels were greater between 10 AM-2 PM in subjects with BD (C). CRH protein levels were consistently lower across the day in subjects with BD compared to control subjects (D). CACNA1C protein levels displayed no fluctuation across the day in subjects with BD (E). Protein values represent pg/ml for SST and CRH and ng/ml for PER2, ANRTL and CACNA1C.

### Associations of Serum Protein Levels with Clinical Measures

Clinical characteristics of BD patients are presented in Supplement Table 2. PER2 serum protein levels were significantly associated with alcohol dependence in subjects with BD after adjusting for age, sex, and venipuncture time (26.1 [95% CI, 12.8, 39.4]; FDR-adjusted p=0.007) (Fig. 4). A similar association with alcohol dependence was observed for SST serum protein levels, which was not significant after false discovery adjustment. Furthermore, associations of SST and PER2 protein levels with lithium response were observed, which did not maintain significance after false discovery adjustment (Fig. 4). No associations of CACNA1C, ARNTL, or CRH protein levels were observed with clinical measures (Suppl. Table 3E).

**Figure 4.**
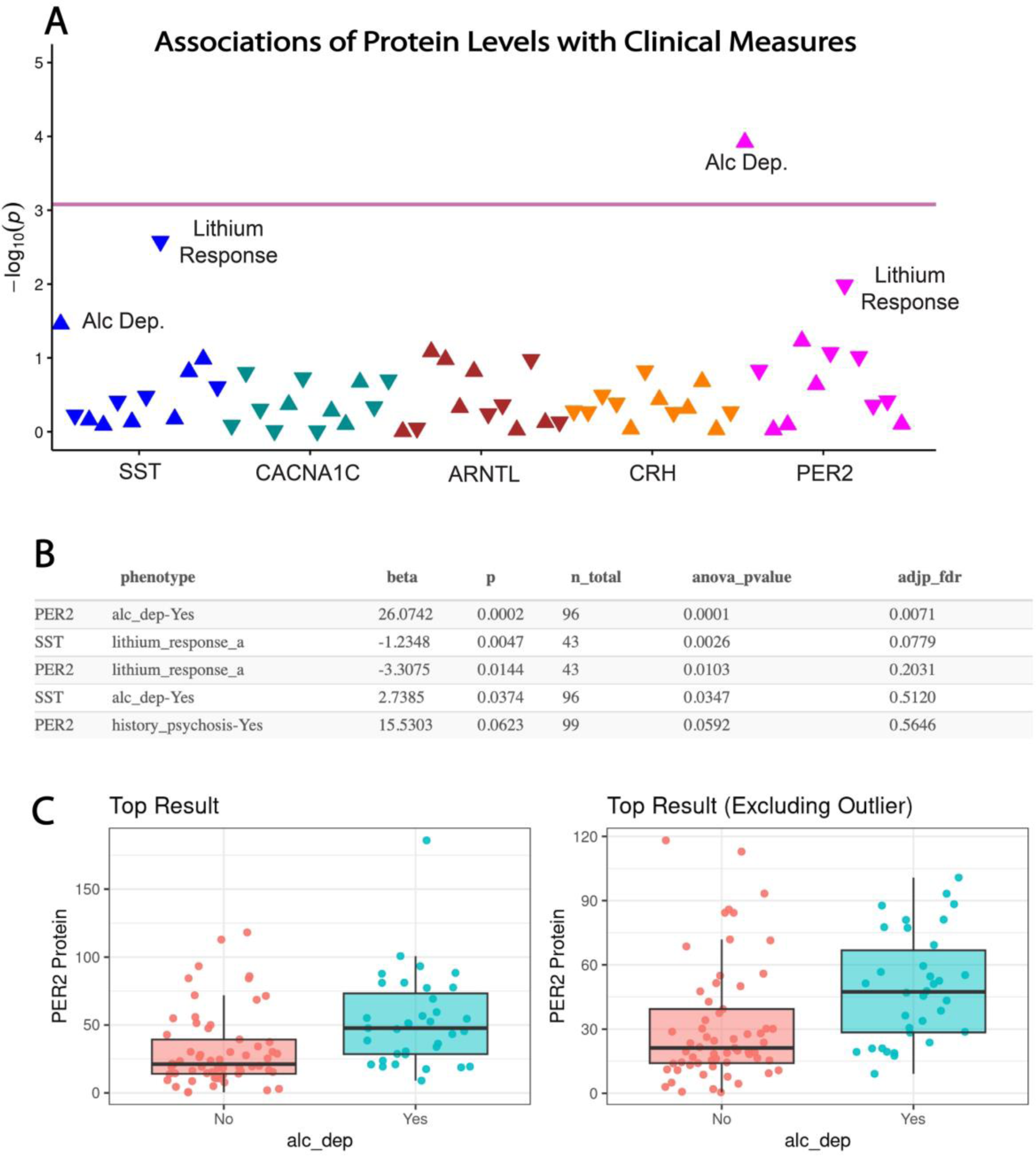
Associations of serum protein levels with clinical measures in subjects with bipolar disorder. (A) PheWAS plot depicting associations of serum protein levels with clinical values. Pink line indicates adjusted P-value significance threshold. PER2 serum levels displayed a significant association with alcohol dependence, whereas associations of PER2 and SST serum levels with lithium response did not maintain significance after FDR correction. (B) Summary table of the top 5 associations with ANOVA p-values and adjusted FDR p-values. (C) Whisker plot depicting the top association of PER2 serum levels with alcohol dependence which maintained significance with the outlier removed. Protein values represent ng/ml for PER2.Lithium response-a represents Alda A score.

## Discussion

Our results represent the first evidence for increased serum CACNA1C protein levels in subjects with BD, as well as the first evidence that CACNA1C protein level is associated with the rs11062170 polymorphism in subjects with BD. Furthermore, increased serum SST levels along with decreased CRH and ARNTL levels suggest disrupted circadian rhythm and stress signaling in the same subjects.

### CACNA1C serum protein and genotype association

Increased CACNA1C serum protein levels in subjects with BD and association of serum CACNA1C level with the rs11062170 risk allele has several implications for BD. CACNA1C polymorphisms are highly implicated in BD by GWAS studies (6–12) and represent promising targets for developing personalized treatments. A recent GWAS also identified enrichment of polygenic risk factors for targets of calcium channel blockers, including the L-type calcium channel blocker isradipine (13). Previous studies have reported increased fMRI activity as well as CACNA1C mRNA expression in the dorsolateral prefrontal cortex of control subjects with two copies of the rs1006737 risk allele (40). Furthermore, increased mRNA expression along with L-type calcium current was reported in cultured induced neurons from people with two copies of the rs1006737 risk allele (41). In contrast, two studies have reported decreased CACNA1C mRNA expression in the superior temporal gyrus and the cerebellum in people with the rs1006737 risk allele (42, 43). These discrepancies in gene expression levels may be due to brain region specific effects including somatic mutations, medication effects or varying representation of more recently reported multiple CACNA1C polymorphisms across these cohorts (44).

As our findings are the first report of protein level associations with the rs11062170 polymorphism of CACNA1C, which was recently identified in a larger GWAS analysis (45), increased levels in patients with two copies of the risk alle may not necessarily reflect prior reported associations of gene or protein levels with other CACNA1C polymorphisms. Furthermore, while several studies have focused on gene polymorphisms and mRNA expression, very few studies have tested associations with protein levels in subjects with BD. While our findings support the hypothesis that CACNA1C protein is increased in the serum of subjects with BD, and is associated with the rs11062170 risk allele, the lack of significant associations of protein levels with clinical measures however suggests that CACNA1C serum levels may not assist in predicting treatment response or clinical symptoms. Future studies with larger cohorts may provide more insight into this question.

### SST and CRH serum protein levels

The most pronounced differences in serum protein levels observed were for SST and CRH. Several studies indicate that decreased brain SST expression is a consistent feature in mood disorders (26, 27). Although this is opposite to the observed increase in serum SST levels in people with BD (Fig.2), increased serum SST levels were reported in rats susceptible to chronic mild stress (29), suggesting that serum levels in BD may be opposite to brain levels, and could represent a potential peripheral marker for predicting susceptibility for depression.

We previously reported altered diurnal rhythm of SST expression in the amygdala of subjects with BD, characterized by lower expression in the morning (28), which coincides with the established increased morning severity of anxiety and depression in people with BD (46–49). In our current study, increased serum SST levels were most pronounced in the morning (Fig. 3) coinciding with the reported morning decrease of SST in the amygdala (28), suggesting a circadian rhythm component to altered SST levels. Altered diurnal rhythms in mice have been reported to increase anxiety in part through SST signaling and in turn to impact CRH expression in the paraventricular nucleus (25). Increased CRH levels may be associated with altered stress response signaling in BD, as indicated by the pronounced decrease in serum CRH levels observed in our study (Fig. 2). Decreased serum CRH in subjects with BD is also in agreement with a prior report using a separate cohort of subjects (30).

The association of serum SST levels with lithium response in subjects with BD (Fig. 4) is in line with previous reports that subjects with mood disorders that were not responsive to lithium had lower SST CSF levels (14–16). Furthermore, these subjects responded to L-type calcium channel blocker treatment (14–16), suggesting that SST may represent a promising biomarker for predicting response to lithium and identifying patients who may benefit from calcium channel blockers. Our findings suggest that serum SST levels may represent a less invasive biomarker for predicting lithium response compared to cerebrospinal fluid SST measures.

CRH levels have also been associated with lithium response. Lithium treatment has been reported to increase CRH levels in the dexamethasone/CRH stress response test, particularly in patients who responded well to lithium treatment (17–19), suggesting that therapeutic effects of lithium are in part through the normalization of hypothalamic stress response signaling and CRH levels.

### ARNTL and PER2 serum protein levels

Decreased serum levels of the core clock molecule ARNTL adds to the extensive evidence for circadian rhythm alterations in people with BD (50–52). ARNTL is a key component of the molecular circadian clock and is the only clock molecule reported to result in loss of rhythmicity in knockout mouse studies (53–55). Decreased levels of ARNTL in subjects with BD were most pronounced in the morning, coinciding with increased SST serum levels (Fig. 3), suggesting that ARNTL and SST alterations may reflect altered diurnal rhythmicity in subjects with BD, as suggested by prior studies (56). Our observations of varying serum protein levels across the day supports the importance of matching cohorts for time of day of serum sample collection. Lack of serum samples collected at night however limits the interpretation of diurnal rhythms in our study. In comparison, we did not detect a significant alteration in serum PER2 levels. It is possible that serum PER2 alterations may be more pronounced in the night which we were not able to analyze in our study, and/or that serum PER2 levels may be less disrupted in subjects with BD compared with ARNTL. Knockdown of ARNTL in the suprachiasmatic nucleus results in depression and anxiety like behaviors in mice (57), suggesting that decreased serum ARNTL levels may reflect a core circadian rhythm disturbance that impacts mood. In addition to its key role in circadian rhythm regulation, ARNTL promotes anti-inflammatory processes (58). Decreased serum levels of ARNTL may contribute to increased peripheral levels of immune system inflammatory processes associated with BD (59–63).

We observed an association of PER2 serum levels with lithium response, which as with a similar association of SST with lithium response, was not significant after false discovery adjustment (Fig. 4). However, this association is also in agreement with studies implicating Per2 rhythmicity alterations in cultured fibroblasts from subjects with BD along with associations of circadian rhythmicity with genetic factors including clock genes and CACNA1C (20, 64). Furthermore, the chronotype of patients as well as the Per2 rhythmicity of their skin fibroblasts predicted responsiveness to lithium therapy (65). Taken together these studies suggest that future studies in larger cohorts with sampling times across the 24-hour cycle that incorporate multiple genetic factors may allow for detection of patients with circadian rhythm dysfunction and lithium non-responders.

The strongest association observed for serum protein levels with clinical measures was for PER2 levels with alcohol dependence in subjects with BD (Fig. 4). Genetic polymorphisms of Per2 have been associated with alcohol use (66–68) and Per2 mutant mice display increased alcohol consumption (66, 69), suggesting Per2 plays a role in alcohol dependence. Clock gene polymorphisms have also been associated with alcohol dependence in subjects with BD (70) suggesting several circadian rhythm genes may represent risk factors for alcohol dependence in this disorder. Furthermore, studies on cultured fibroblasts from patients with alcohol use disorder (AUD) demonstrated that the period of Per2 expression rhythms was inversely correlated with severity of AUD (71). In addition to clock gene polymorphisms, the CACNA1C rs1034936 genotype has been associated with alcohol dependence in people with BD (37), and increased CACNA1C expression has been reported in a preclinical model of alcohol dependence (72).

### Technical Limitations

Although the cohort in our current study represented a large sample size in comparison to other studies on serum protein markers in BD (30, 59–62), our sample size was limited for analysis of genetic associations with serum levels and clinical measures. Moreover, multiple genetic factors are likely to contribute to altered protein levels and clinical measures. Future studies with larger cohorts will provide insight into polygenic risk factors associated with serum levels and clinical measures in subjects with BD. Furthermore, the inability to collect serum samples during the night limits our ability to interpret diurnal rhythms in serum protein levels and potential circadian rhythm alterations. Serum level collections from participants in sleep lab settings may allow for development of serum sample cohorts for improved studies on serum diurnal rhythm levels in BD.

### Summary

In summary, our results represent the first evidence for increased levels of CACNA1C in the serum of subjects with BD, associated with two copies of the rs11062170 risk allele. Together with alterations in SST, ARNTL, and CRH, these findings suggest peripheral markers of circadian rhythm regulation and stress response are altered in the same cohort of subjects with increased CACNA1C levels in subjects with BD. Our results also add to the growing evidence for the role of PER2 in alcohol dependence in subjects with BD. Future studies in larger cohorts may assist with identifying serum markers that predict response to lithium therapy and identify patients who may benefit from chronotherapy or calcium channel blockers.

## Supporting information

Allen IV et al Suppl Materials

## Acknowledgements

We acknowledge the Mayo Clinic Biobank (MCB) research team, as well as the patient-participants who consented to participate in this research program. We also acknowledge the Mayo Clinic Center for Individualized Medicine for support of the Mayo Clinic Biobank, and Regeneron Genetics Center for providing genetic data for participants which was utilized for this project for our candidate SNP. Establishment of the Bipolar Disorder Biobank was supported by a generous gift from the Marriot Family and the Mayo Clinic Center for Individualized Medicine.

## Data Availability Statement

The data and original contributions presented in this study are included in the manuscript or supplemental materials. Further inquiries can be directed to the corresponding author.

