## Supplementary material for "Differential Serum Levels of CACNA1C, Circadian Rhythm and Stress Response Molecules in Subjects with Bipolar Disorder: Associations with Genetic and Clinical Factors": Allen IV et al Suppl Materials

**Supp Table 1.** Protein levels

| Characteristics | BD (N=100) | Control (N=100) | Difference (Mean 95% CI) | P |
|---|---|---|---|---|
| | N (%) or Mean (SD) [Median (IQR)] | | | |
| Age | 46.9 (13.1) | 46.7 (13.1) | 0.179 (-0.280, 0.637) | 0.442 |
| Gender | | | | NA |
| Female | 62 (62%) | 62 (62%) | NA | |
| Male | 38 (38%) | 38 (38%) | NA | |
| Venipuncture time, h | 12.1 (2.1) | 12.1 (2.1) | 0.000 (-0.001, 0.001) | 0.783 |
| Proteins | | | | |
| SST[a] | 25.1 (5.8) [23.5 (22.9, 25.7)] | 21.5 (7.5) [23.9 (19.7, 25.5)] | 3.6 (1.7, 5.5) | <0.001 |
| CACNA1C[b] | 0.395 (0.011) [0.392 (0.388, 0.396)] | 0.363 (0.094) [0.389 (0.354, 0.399)] | 0.032 (0.013, 0.051) | 0.001 |
| ARNTL[b] | 0.528 (0.068) [0.499 (0.480, 0.597)] | 0.555 (0.092) [0.510 (0.487, 0.644)] | -0.027 (-0.051, -0.003) | 0.029 |
| CRH[b] | 6.24 (1.54) [6.58 (5.09, 6.89)] | 8.65 (1.36) [8.56 (7.81, 9.38)] | -2.41 (-2.79, -2.04) | <0.001 |
| PER2[d] | 38.0 (30.5) [28.4 (17.2, 53.6)] | 31.3 (23.7) [25.2 (14.3, 42.8)] | 6.7 (-1.4, 14.7) | 0.103 |

p-values calculated from paired t-tests.
[a]N=96 pairs because missing data from n=4 pairs.
[b]N=98 pairs because missing data from n=2 pairs.
[d]N=95 pairs because missing data from n=5 pairs.

**Supp Table 2.** Clinical characteristics

| Characteristics | BD (N=100) N (%) or Median (IQR) |
|---|---|
| Diagnosis[a] | |
| BD I | 58 (62%) |
| BD II | 36 (38%) |
| Alcohol dependence[b] | 36 (37%) |
| Hx psychosis | 19 (19%) |
| Hx suicide attempt | 38 (38%) |
| Current episode[c] | |
| Manic or hypomanic | 11 (13%) |
| Major depressive | 45 (53%) |
| Euthymic | 29 (34%) |

| Characteristics | BD (N=100)<br>N (%) or Median (IQR) |
|---|---|
| Rapid cycling[d] | 36 (67%) |
| CGI Scale[e] | |
| Mania | 2.0 (1.0, 3.0) |
| Depression | 3.0 (1.0, 4.0) |
| Chronotype[f] | |
| Non evening type | 65 (68%) |
| An evening type | 30 (32%) |
| Insomnia severity[g] | 2.0 (2.0, 3.0) |
| Lithium (current) | 23 (23%) |
| Lithium Alda A score[h] | 6.0 (3.5, 7.0) |

Abbreviations: BD, bipolar disorder; CGI, Clinical Global Impressions

[a]N=94 because missing data from n=6.
[b]N=97 because missing data from n=3.
[c]N=85 because missing data from n=15.
[d]N=54 because missing data from n=46.
[e]N=96 because missing data from n=4.
[f]N=95 because missing data from n=5.
[g]N=51 because missing data from n=49. Insomnia severity is calculated as the maximum insomnia severity score [0-3] across sleep onset, mid-nocturnal, and early morning.
[h]N=43 because missing data from n=57.
